# Genetic and Health Determinants of Cancer Risk in Bangladeshi and Pakistani South Asians in the UK

**DOI:** 10.64898/2025.12.09.25341887

**Authors:** Abu Zafer Mohammed Dayem Ullah, Ashitha Joby, Graeme John Thorn, Lewis George Emin James, Isabelle Sanders, Genes & Health Research Team, Claude Chelala

## Abstract

South Asian populations remain underrepresented in cancer genomics, despite elevated risk for certain malignancies and distinct clinical profiles. This gap is especially pronounced for British Bangladeshi and Pakistani communities. We analysed data from 57,416 individuals of Bangladeshi and Pakistani ancestry in the UK-based *Genes & Health* cohort, integrating electronic health records, cancer registry data, and whole-exome sequencing (n=43,462). Among them, 2,782 (4.8%) had cancer, with earlier onset across multiple types compared to national benchmarks. A phenome-wide case–control analysis identified 147 significant cancer–comorbidity associations, with larger effect sizes observed for topographically concordant cancer-comorbidity pairs, and stronger associations for systemic disorders such as obesity and hypertension. Exome-wide analyses revealed 39 variant-level and 31 gene-level associations, over 60% novel and enriched for rare coding variations. Notable loci included *ARHGAP45*, *CBR1*, *FKBP6*, *NF1*, and *ZNF155*, with several ancestry-specific associations. These findings highlight distinct cancer risk architectures in this South Asian subpopulation, emphasising the need for population-tailored risk models and screening strategies.

## Main

Cancer continues to be one of the leading global health challenges, with an estimated 20 million new cases and 10 million deaths in 2022 alone.^1^ Despite considerable advances in prevention, early detection, and treatment, substantial disparities persist in cancer incidence, presentation, and survival outcomes across population groups identified by geographical region, ethnicity or income level.^1,2^ People of South Asian ancestry, who comprise nearly a quarter of the global population and are one of the fastest-growing minority groups in Western countries — including the United Kingdom (8.5%), Canada (7.1%), Australia (6.6%), and the United States (1.5%) — remain severely underrepresented in healthcare research and genomic studies.^3–6^

Historically, South Asian migrants to Western nations have exhibited lower overall cancer incidence compared to their host populations, particularly for malignancies such as breast, lung, colorectal, and prostate cancer — diseases that collectively account for a substantial proportion of cancer morbidity and mortality in Western countries.^7–9^ However, growing evidence indicates a rapid convergence in cancer incidence rates between South Asians and their host populations, especially with increasing age and length of residence.^10^ Furthermore, South Asian patients are disproportionately diagnosed at younger ages or with more advanced-stage disease compared to their White counterparts,^9,11,12^ a disparity often attributed to lower participation in routine cancer screening programs.^13–15^ While socio-cultural barriers and health care system factors contribute to these disparities, such reasoning may not fully account for the observed differences in cancers that lack standardised population-based screening protocols.

South Asians experience a disproportionately high burden of chronic health conditions, including type 2 diabetes mellitus, metabolic syndrome, central obesity, and chronic viral hepatitis.^16–22^ These conditions are well-established risk factors for several malignancies — notably liver, colorectal, pancreatic, and endometrial cancers — and are thought to have a strong inherited genetic component in South Asian populations.^23,24^ However, our understanding of the genetic basis of complex diseases, including cancer, remains largely biased towards populations of European ancestry.^25^ To date, individuals of South Asian descent have constituted only about 2% of participants in published genome-wide association studies (GWAS).^26^ Large, diverse resources such as the UK Biobank (∼1%), the All of Us Research Program (∼1%), and the Mass General Brigham Biobank (<1%) still include insufficient South Asian representation, limiting the ability to conduct meaningful genetic analyses of cancer risk.^27^ This persistent underrepresentation significantly limits our ability to fully understand cancer biology, improve risk prediction, and develop personalised therapeutic strategies for this growing diverse population.

Understanding the intersection between clinical risk factors and genetic predisposition in South Asian populations can uncover valuable insights for cancer prevention and management tailored to this group. Such understanding is crucial for developing culturally relevant prevention strategies, enhancing early detection, and creating risk models specific to this population. To advance this objective, the present study uses the *Genes & Health* (G&H) cohort — the largest longitudinal genotype-phenotype resource of Bangladeshi and Pakistani individuals recruited in the United Kingdom. This cohort included approximately 57,000 participants,^28^ with around 2,800 cancer cases across 33 cancer types encompassing both primary sites and broader organ systems.

We conducted a comprehensive characterisation of cancer phenotypes and associated clinical risk factors within this cohort, leveraging extensive electronic health records and cancer registry data. In addition, we explored the contribution of rare and low-frequency protein-coding genetic variants in cancer susceptibility by using whole-exome sequencing data from a subset of approximately 43,000 individuals. These genetic associations were evaluated in the context of existing GWAS studies, biomarker and drug target databases. Finally, we assessed whether our ancestry-specific findings were consistent with patterns observed in the cancer cohort of *Genomics England* (GEL) 100,000 Genomes Project (100K GP).^29^

This study aims to fill critical gaps in the current literature by elucidating the combined effects of clinical and inherited risk factors for cancer in an underrepresented ethnic group. The insights gained will serve as a foundation for population-specific biomarker discovery, and will inform strategies for improving risk stratification, refining screening protocols, and developing personalised therapeutic approaches for South Asian communities in the Western world.

## Results

### Cancer incidence and age at diagnosis

The G&H cohort included 57,416 South Asian individuals of Bangladeshi and Pakistani origin (SAS-BP), of whom 2,782 (4.85%) had a cancer diagnosis recorded in their electronic health records (EHR) (Fig. 1) A high, degree of concordance (99%) was observed between EHR-coded ethnicity and genetic Ancestry (gAncestry) derived from genotyping array data of 51,166 participants (Supplementary Fig. 1), which aligns with the recruitment criteria of this cohort. After adjusting for gAncestry data (Methods), the study cohort comprised 52% Bangladeshi and 48% Pakistani origin participants (Supplementary Table 1).

**Figure 1.**
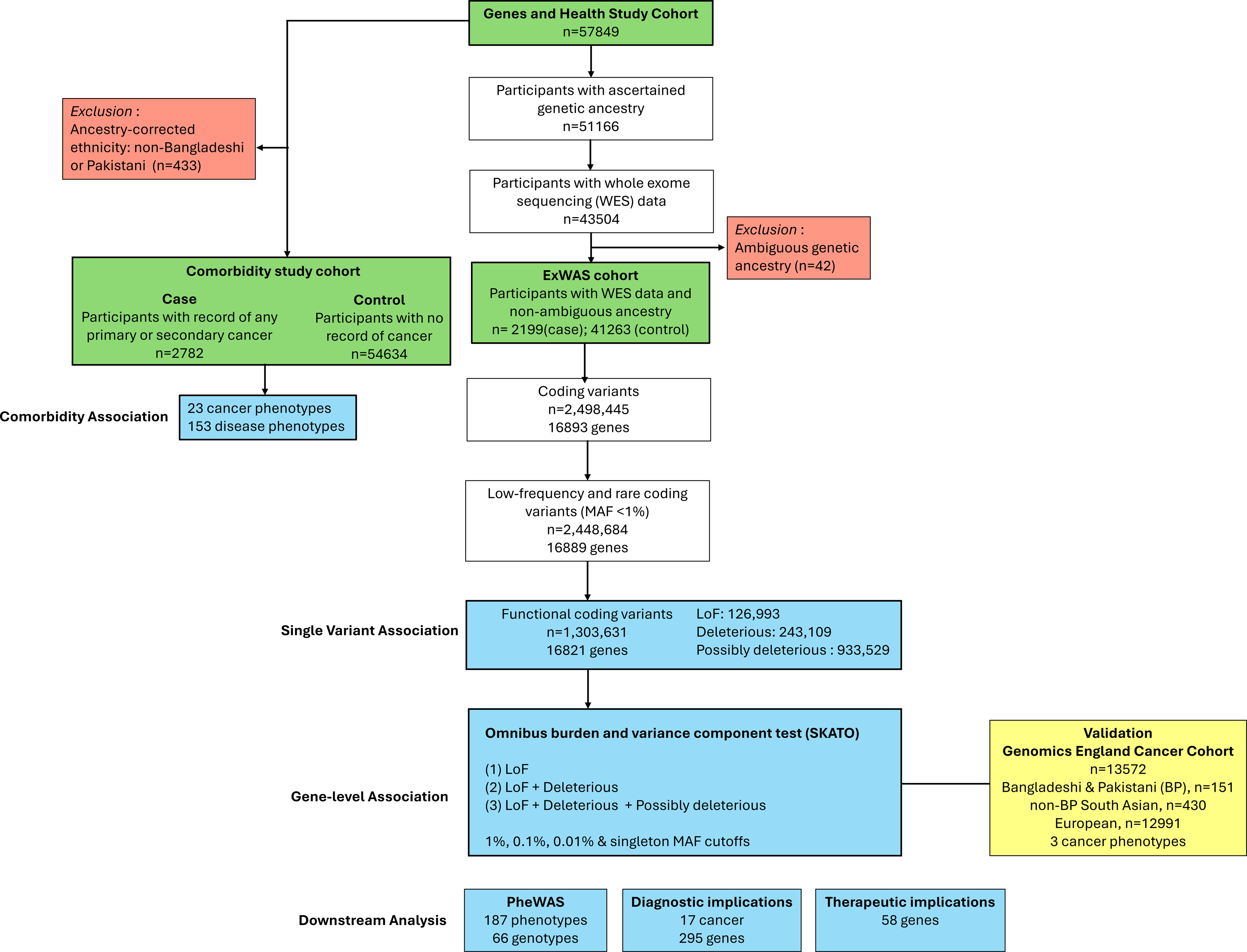
Study overview for germline variants and clinical determinants exploration across cancer phenotypes in British Bangladeshi and Pakistani individuals. Two streams of analysis were conducted on the Genes & Health study cohort (n=57,416). Comorbidity association stream focused on exploring associations between cancer phenotypes and pre-existing non-cancer medical conditions. Exome-wide association study (ExWAS) stream focused on discovering associations between cancers and low-frequency loss-of-function and missense coding variants, at single variant as well as gene level, using a subset of participants with whole exome-sequenced data (n=43,462). Significantly associated genes and/or variants were further tested for differential germline profile in Genomics England Cancer Cohort (n=13,572), association with other phenotypes, and viability in germline testing and known treatment regimens.

Cancers originating from the gynaecological, digestive, and hematopoietic systems were more prevalent, with breast, prostate, cervical and colorectal cancers representing the most prevalent primary sites (Supplementary Table 2). The prevalence of 13 distinct primary and 10 aggregate cancer types within the cohort stratified by demographic characteristics are shown in Figure 2. Excluding gender-specific cancers, the overall incidence of cancer was generally higher in men than women. However, the prevalence of (female) breast cancer was notably high in this population (1.3%) which skewed the overall cancer statistics towards women.

**Figure 2.**
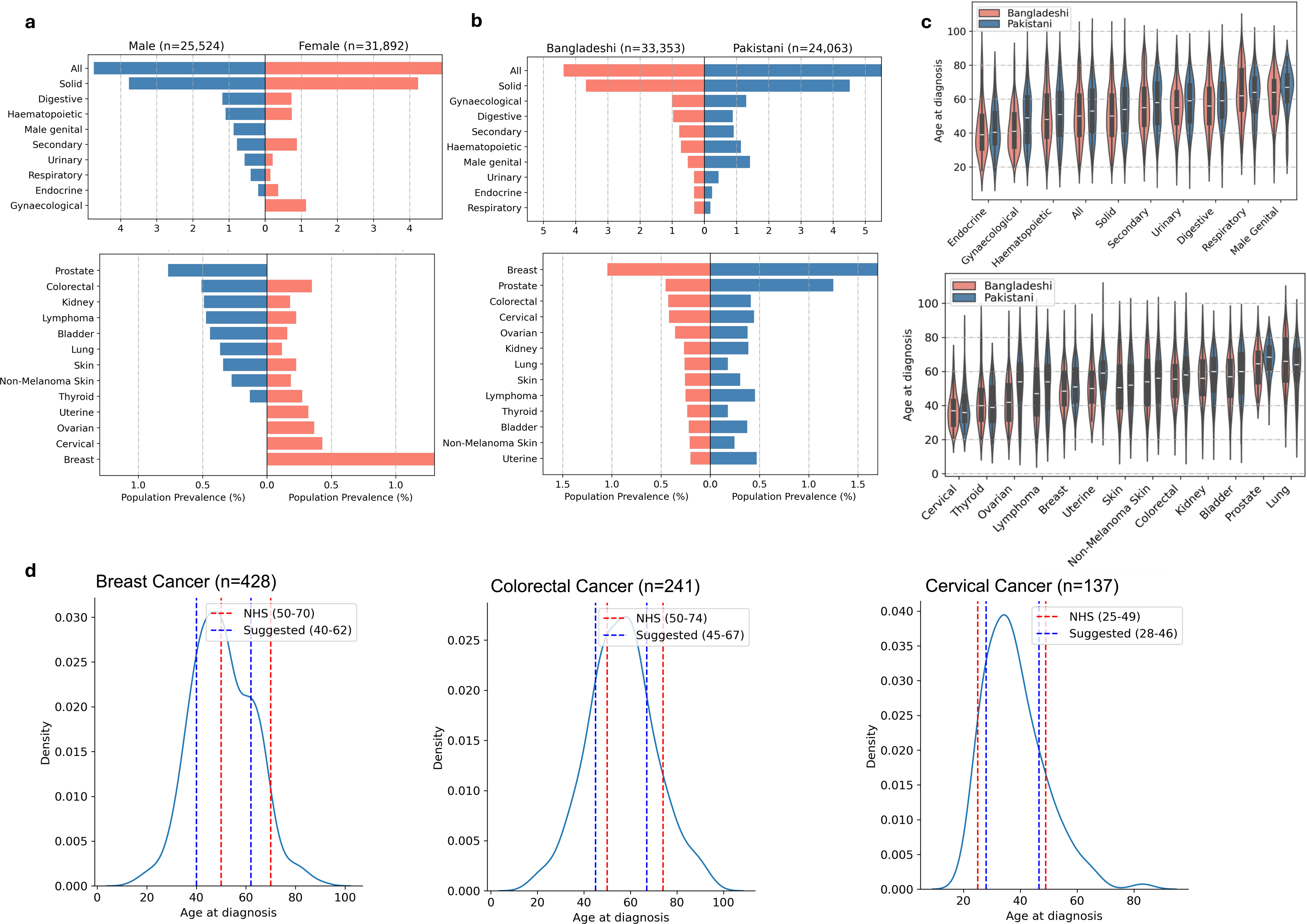
Demographic features of cancer patients within the Genes & Health study cohort. **a-b,** Bi-directional barplots showing prevalence of different cancers within the study cohort, stratified by sex and ethnicity. Each bar represents the prevalence of a cancer in the specific demographic subgroup - male, female, Bangladeshi, Pakistani (n=24,063). EHR-coded ethnicity of participants are corrected for genetic ancestry determined from genotyping array data, where available. Barplots are displayed in descending order of prevalence in male and Bangladeshi subgroups (from top to bottom). **c,** Grouped violin plots with overlaid boxplots showing the distribution of age of cancer diagnosis, stratified by genetic ancestry corrected ethnicity. Plots are displayed in ascending order of median diagnosis age for cancers in Bangladeshi subgroup (from left to right). **a-c,** Each pair of plots show demographic distribution for cancers of 13 primary sites (bottom) and 10 aggregate types (top). The number of participants in each cancer are: 2782 (all), 2312 (solid), 534 (digestive), 515 (haematopoietic), 479 (secondary),428 (breast), 361 (gynaecological), 241 (colorectal), 222 (male genital), 212 (urinary), 196 (prostate), 193 (lymphoma), 182 (kidney), 164 (endocrine), 163 (bladder), 159 (skin), 149 (respiratory), 137 (cervical), 130 (lung), 129 (non-melanoma skin), 121 (thyroid), 117 (ovarian) and 102 (uterine). **d,** Visualisation of the age-based screening windows for breast, colorectal and cervical cancers in the British Bangladeshi and Pakistani populations. The red vertical lines show current screening windows deployed in the UK National Health Service. The blue vertical lines show the suggested windows as dictated by this study.

When comparing Bangladeshi and Pakistanis participants, the latter exhibited a higher cancer diagnosis rate, with the disparity being particularly pronounced for breast, reproductive organs and lymphoma cancers. However Bangladeshi participants were diagnosed at a younger age for these cancer types (Breast: 48 years vs 51 years; Ovarian: 42 years vs 54 years; Uterine: 50 years vs 59 years; Prostate: 64.5 years vs 68.5 years; Lymphoma: 46 years vs 54 years). Cervical cancer patients were the youngest group, with a median age of 36 years (IQR:13 years), while prostate cancer patients were the oldest with a median age of 67 years (IQR:14 years).

Approximately, half of the cancer patients in our cohort were diagnosed by the age of 51 (IQR:24 years) (Fig. 2c), which is notably earlier than the broader, predominantly White population in the country. In the general UK population, only 14% cancer patients are diagnosed by the age of 50.^30^ This early onset was also evident in GEL cohort (Supplementary Table 3), where median age of cancer diagnosis was 64 years (IQR: 18 years) for Europeans and 57 years (IQ: 19 years) for South Asians outside of Bangladeshi or Pakistani ancestry (SAS-nBP; presumably largely Indian ancestry). This earlier age of diagnosis was particularly pronounced for breast, ovarian, uterine, colorectal, and respiratory cancers. To assess whether the current UK NHS screening guidelines are equitable for this cohort,^31^ we further stratified the breast, colorectal and cervical cancer groups based on the central 60% of the age distribution (Fig. 2d). We identified cancer-specific intervals: 40-62 years for breast cancer, 45-67 years for colorectal cancer, and 28-46 years for cervical cancer. These findings suggest that bringing forward the screening windows for breast and colorectal cancers by approximately 10 and 5 years, respectively, would provide equitable benefit for the SAS-BP population. Interestingly, a narrower screening window for cervical cancer would yield similar benefits.

### Comorbidities associated with cancer

To explore pre-existing non-malignant conditions that might contribute to or reflect cancer susceptibility, we conducted a case-control study to examine for associations between 23 cancer types and 151 phenotypes. Comorbidities were defined using a validated phecode framework, which mapped diagnostic codes from both primary care and secondary care into clinically meaningful categories (Methods). A total of 3488 tests were performed. After applying Bonferroni correction for multiple testing with a significance threshold of P≤1.5×10^−5^, we identified 157 significant associations for aggregate cancer types and 32 for primary cancer types. Most of these significant associations were observed in the top five aggregate cancer types (Fig. 3a). Several cancer pairs exhibited high degree of correlation (Pearson’s r>0.8), including all–solid, endocrine–thyroid, respiratory–lung, prostate–male genital, skin–non-melanoma skin, bladder–urinary, and kidney–urinary cancers. In cases where common signal emerged from these pairs, we retained the association with the lower *P value*, resulting in 147 unique cancer-phenotype associations (Supplementary Table 4).

**Fig. 3:**
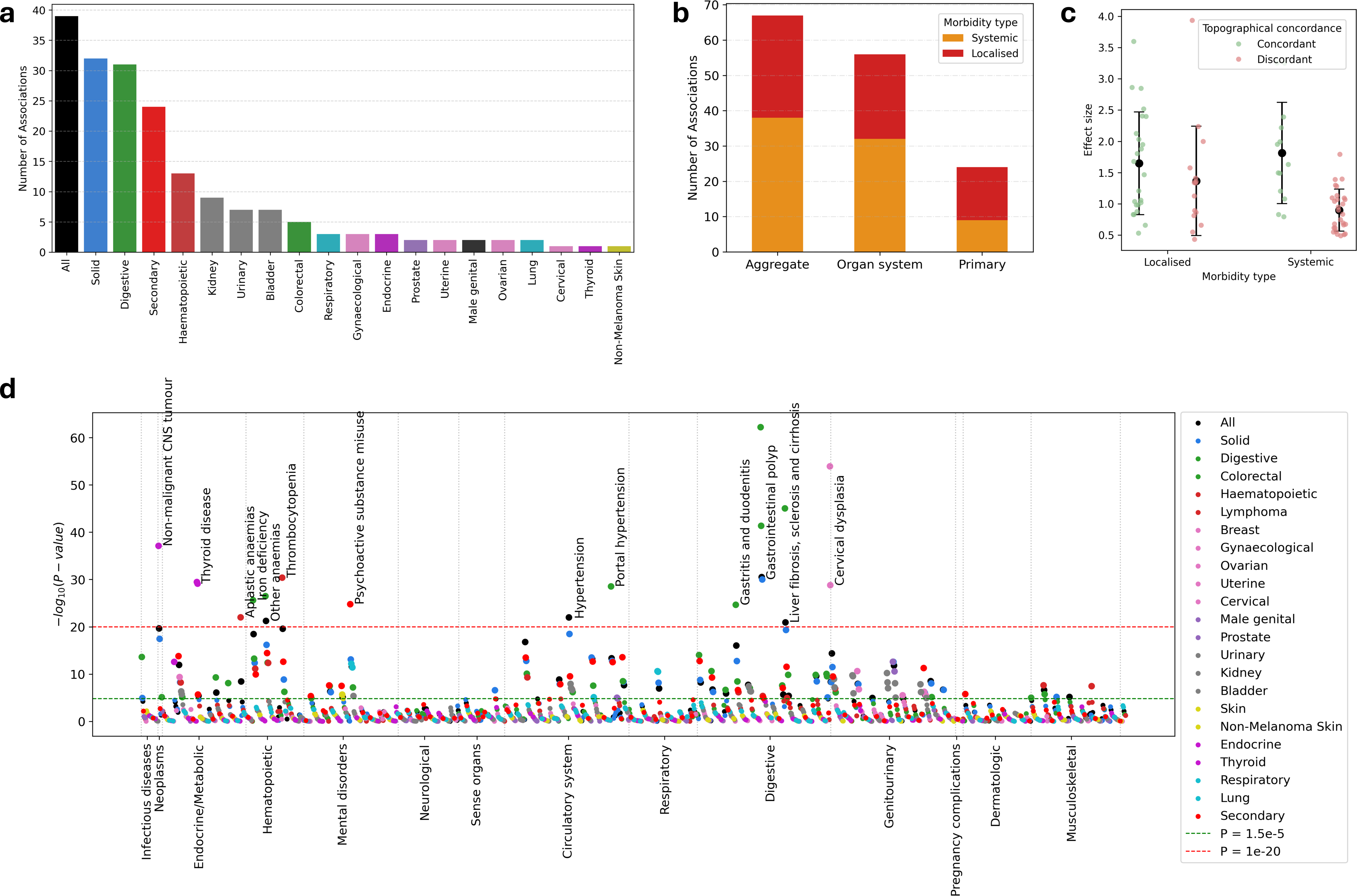
Summary of comorbidity association study results. **a,** Barplot showing number of significant associations found between each cancer type and pre-existing medical conditions, **b,** Distribution of systemic and localised pre-existing conditions significantly associated with cancers, grouped by topographical distribution (Primary – cancers in a single organ; Ogran system – cancers in multiple primary sites from same organ system; Aggregate - *all*, *solid* and *secondary* cancers with heterogeneous origins) **c,** Strip plot with mean ± standard deviation showing effect size of associations, grouped by types of medical conditions: systemic (n=41) vs localised (n=39) and topographical alignment: concordant (n=37) vs discordant (n=43). The y□axis shows the log-scaled odds-ratio with each dot representing an association. Associations with *all*, *solid* and *secondary* cancers were excluded. **d,** Manhattan plot showing associations (n=147) between 23 cancers and pre-existing medical conditions stratified by 14 phenotype groups. The green dashed line represents the significance threshold at P□=□1.5×10^-5^ to account for multiple hypothesis testing. Points (associations) are color coded according to cancer type. Only pre-existing conditions from highly significant associations (above red line; P□≤1×10^-20^) are labelled (n=20).

The top five significant associations identified were gastrointestinal polyp or chronic liver disease prior to digestive cancer (P=6.2×10^−63^, P=9.2×10^−46^), cervical dysplasia prior to cervical cancer (P=1.1×10^−54^), thrombocytopenia (low platelet count) in haematological cancer (P=4.1×10^−31^), and benign tumour in the central nervous system (CNS) prior to endocrine cancer (P=7.7×10^−38^) - the latter showing the largest effect size (OR: 51.1, 95% CI: 28.1-93.1). Among systemic disorders, obesity, hypertension, chronic renal disease and anaemias accounted for 40% (29 of 73) of the associations. Notably, hypertension and obesity, two highly prevalent conditions in the community (26% and 21%, respectively), were associated with increased odds of digestive, haematological and renal cancers.

Across the cancer spectrum, the highest number of pre-existing conditions was observed for digestive system diseases/disorders (25%), followed by genitourinary system conditions (20%) and circulatory system diseases (16%) (Fig. 3d). After excluding associations related to three aggregate cancers encompassing heterogeneous sites (*all cancers combined*, *solid* cancers and *secondary* cancers), 37 of the remaining 80 associations (46%) represented topographically concordant pre-existing conditions with stronger effect sizes. This highlights the impact of exposure to localised disorders on carcinogenesis within the same organ system (Fig. 3b-c). In contrast, systemic discordant conditions displayed weaker associations across cancer sites.

Most associations with systemic disorders align with known risk factors for the corresponding cancers. One notable finding was the association between fibromyalgia with haematopoietic (OR: 3.7 [2.3-5.8]; P=2.4×10^−8^) and digestive cancers (OR: 2.9 [1.9-4.5]; P=1.9×10^−6^). While fibromyalgia, a chronic pain disorder, shares overlapping symptoms with cancer, such as pain and fatigue, no direct link has been established. Our findings may reflect a misdiagnosis of cancer as fibromyalgia, a common issue due to the crossover in symptoms.^32^ We also observed a strong association between substance misuse and respiratory cancer (OR: 4.0 [2.8-5.9]; P=6.5×10^−13^), as well as with digestive and bladder cancers.

Subgroup analysis by sex and ethnicity identified 55 significant associations, of which 42 (76%) were linked to either male or Bangladeshi participants (Supplementary Table 5). These results suggest an opportunity to expand surveillance efforts for these groups. Specific associations of interest included: Bangladeshi participants – gastritis/duodenitis in digestive cancer (OR: 3.3 [2.6-4.2]; P=2.4×10^−23^), peripheral arterial disease in bladder cancer (OR: 8.7 [4.0-18.7]; P=3.0×10^−8^), polymyalgia rheumatica in haematopoietic cancer (OR: 11.1 [4.3-29.0]; P=8.1×10^−7^); SAS-BP men – iron deficiency in digestive cancer (OR: 5.0 [3.8-6.7]; P=1.4×10^−27^), COPD in respiratory cancer (OR: 4.8 [3.0-7.7)]; P=3.7×10^−11^); SAS-BP women – cerebrovascular disease in endocrine cancer (OR: 6.7 [3.1-14.6]; P=1.6×10^−6^)

### Coding variant distributions

We analysed whole-exome sequencing (WES) data from 43,462 individuals within G&H, following rigorous quality control procedures and excluding participants with ambiguous gAncestry (Methods). The cohort included 2,199 cancer patients (5.1%) and 41,263 cancer-free participants. 54% of the participants were of Bangladeshi gAncestry, which closely mirrors the composition of the overall study population. We identified 2.5 million autosomal variants within the coding regions of genes (Table 1), of which 98% were classified as low-frequency variants (minor allele frequency (MAF) ≤1% across the cohort; Supplementary Table 6). This catalogue represents a 4.2-fold increase over the number of South Asian participants in the UK Biobank dataset, with 1.4-fold increase in the number of coding variations.^33^

**Table 1.** Coding variants discovered in exome sequencing data from the Genes & Health WES cohort.

|  | Genes & Health (n=43,462) |  | Variants not observed in gnomAD 4.1 Exomes |  |
| --- | --- | --- | --- | --- |
| Variant category | Number of variants, N (% with MAC $\geq 1$) | Variants per participant, median (IQR) | South Asian (n=43,129) N (%) | All (n=730,947) N (%) |
| Coding regions | 2,498,445 (42.76) | 18,373 (305) | 1,155,378 (46.24) | 695,288 (27.83) |
| **Predicted function** |  |  |  |  |
| LoF | 127,662 (53.76) | 191 (15) | 81,570 (63.90) | 61,940 (48.52) |
| Start lost | 3,033 (49.65) | 7 (2) | 1,743 (57.47) | 995 (32.81) |
| Stop gained | 43,145 (52.23) | 52 (8) | 25,054 (58.07) | 15,808 (36.64) |
| Stop lost | 1,319 (51.32) | 13 (3) | 777 (58.91) | 606 (45.94) |
| Splice donor | 14,203 (53.47) | 13 (4) | 9,032 (63.59) | 6,404 (45.09) |
| Frameshift | 55,489 (54.97) | 91 (10) | 37,993 (68.47) | 32,644 (58.83) |
| Splice acceptor | 10,473 (55.56) | 14 (3) | 6,971 (66.56) | 5,483 (52.35) |
| Missense | 1,553,669 (43.73) | 8,354 (163) | 734,794 (47.29) | 441,008 (28.38) |
| Deleterious | 243,109 (48.08) | 79 (11) | 125,796 (51.74) | 74,776 (30.76) |
| Possibly deleterious | 933,529 (44.23) | 2,138 (66) | 447,120 (47.90) | 271,557 (29.09) |
| Likely benign | 377,031 (39.70) | 6,132 (117) | 161,878 (42.93) | 94,675 (25.11) |
| In-frame indels | 31,711 (43.26) | 199 (13) | 17,082 (53.87) | 12,411 (39.14) |
| Insertion | 8,742 (41.85) | 91 (9) | 4,997 (57.16) | 4,108 (46.99) |
| Deletion | 22,969 (43.79) | 108 (10) | 12,085 (52.61) | 8,3030 (36.15) |
| Synonymous | 785,369 (39.02) | 9,621 (161) | 321,915 (40.99) | 179,914 (22.91) |
| Other coding | 34 (38.23) | 1 (0) | 17 (50.00) | 15 (44.12) |

Variant functions are predicted based on mapping on the canonical transcript MAC, minor allele count; IQR, interquartile range (quartile 3 - quartile 1).

Among the variants identified, 1,553,669 were missense (median of 8,354 per individual), and 127,662 were putative loss-of-function (LoF) variants (median of 191 per individual) (Table 1). Of the missense variants, approximately 16% (243,109) were predicted to be deleterious by 5 prediction algorithms (henceforth referred to as ‘deleterious missense’ variants) while 60% (933,529) were predicted to be deleterious by at least one algorithm (henceforth referred to as ‘possibly deleterious missense’ variants). Notably, approximately 43% of variants were observed only once in the dataset (singleton variants; Supplementary Fig. 2).

While variant distributions showed little to no difference in between cancer and non-cancer groups across variant types, significant differences were observed between Bangladeshi and Pakistani participants, with a higher number of variants identified in the Bangladeshi group (median: 18,451 vs 18,204; Supplementary Table 7). Almost half of these variants were not reported in gnomAD v4.1 South Asian Exomes, and a quarter were not reported in any ancestry group (Table 1). The novelty was even more pronounced for LoF variants, with 64% unseen in South Asian populations and 49% overall. This distinctive catalogue of coding variants, coupled with the relatively large sample size, provides a unique opportunity to investigate germline genetics in cancer within this under-researched ethnic group.

### Coding variant associations

We conducted a variant-level exome-wide association study (ExWAS) to investigate associations between 22 cancer phenotypes (excluding secondary cancers) and low-frequency/rare coding variants (0.005%< MAF≤1%), observed in at least five participants, resulting in 6.2 million individual tests. The smallest cancer cohort, uterine cancer, included 76 cases. The analysis focused on 331,291 LoF variants (including stop gain, stop lost, start lost, frameshift and essential splice variants), as well as deleterious and possibly deleterious missense variants across 16,422 protein-coding genes.

The G&H cohort is characterised by a high rate of kinship, with 36.5% of participants being first cousins or closer.^34^ The rate was even higher among the cancer cases, with 61.5% having at least one second-degree or closer relative (Supplementary Table 1). Given this familial structure, association analyses were performed using the whole-genome regression approach implemented in REGENIE, which accounts for relatedness, population structure and polygenicity. This approach also employs a fast, approximate Firth regression approach for binary outcomes.

Using a P value threshold of 3×10^-6^ (Methods), we identified 47 significant associations (Figure 4a; Supplementary Table 8). In cases where a common signal was observed from highly correlated cancer pairs, we retained the association with the lower P value, resulting in 39 unique cancer-variant associations. Of these, 38 were derived from rare variants (MAF<0.1%). 28% (11 of 39) of these variants had not been previously reported in South Asian populations. The rarest significant variant was a deleterious missense variant in the *Carbonyl Reductase 1* gene (CBR1:*p*.Arg74Pro), which plays a critical role in drug metabolism and was associated with aggregated solid cancers (cohort MAF of 0.006%). All significant associations exhibited a consistent positive direction of effect, contrasting with disease-associated non-coding variants, which can variably affect gene expression.^35^ The largest effect was observed for a frameshift variant in the *Zinc Finger Protein 155* (ZNF155) gene, which was associated with prostate cancer (log-OR: 7.0 [5.0-9.1], P=2.8×10^-7^) – the only significant LoF variant association.

**Fig. 4:**
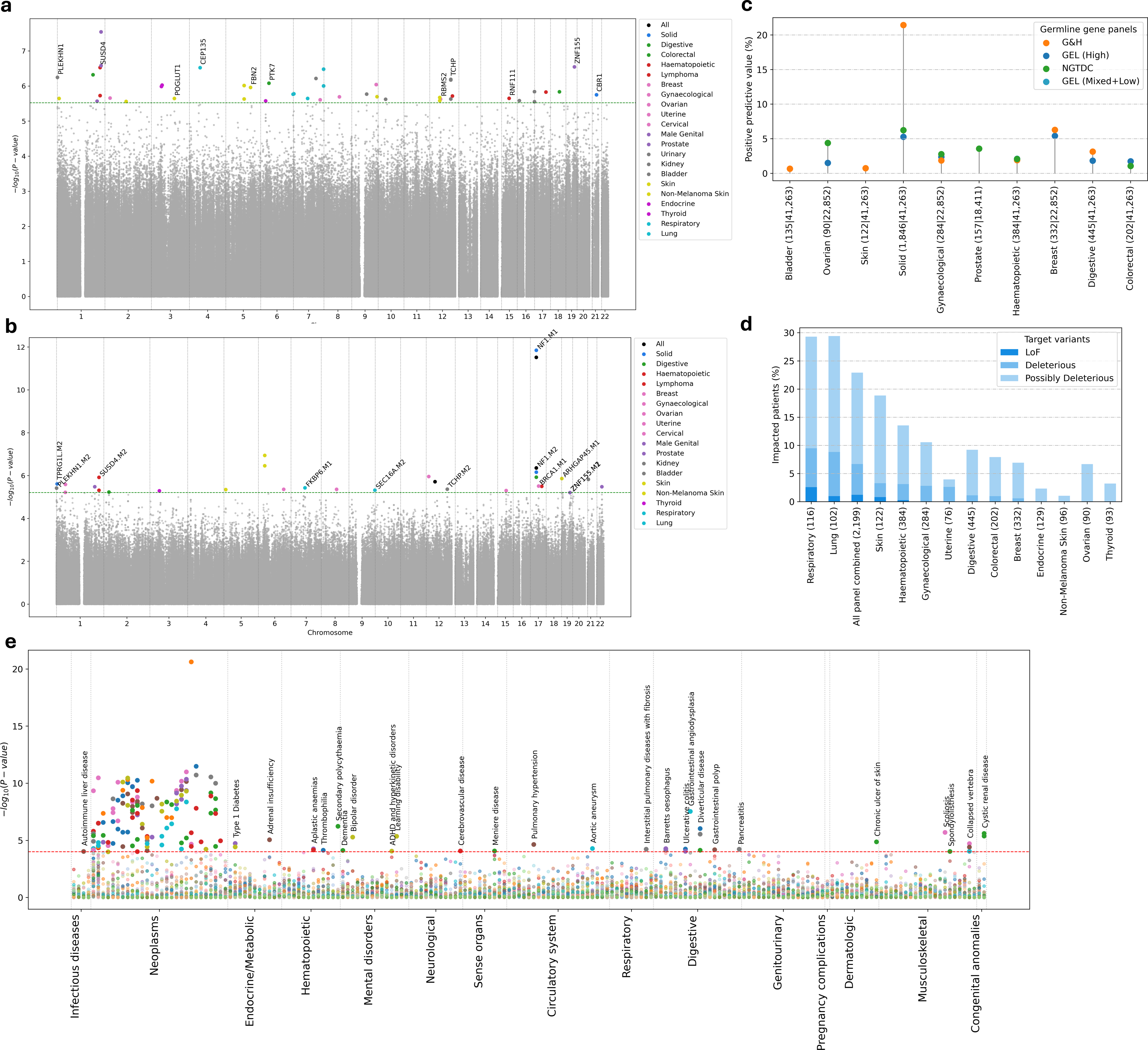
Summary of exome-wide association study results and downstream analyses. **a-b,** Multi-trait Manhattan plot exhibiting the landscape of **(a)** variant–cancer and **(b)** gene-cancer associations across the 331,291 LoF, deleterious and possibly deleterious missense coding variants, with the −log_10_ of the *P*□value on the *y*□axis. The green dashed line represents study-specific significance threshold at **(a)**P=3×10^-6^ and **(b)** P=6.2×10^-6^ to account for multiple hypothesis testing. Each point (association) above the line (significance threshold) is color coded according to cancer phenotype. **a.** Only genes representing LoF or deleterious missense variants are labelled. **b.** A maximum of 3 masks per gene was tested representing aggregation of (M1) LoF, (M2) LoF + deleterious missense, (M3) LoF + deleterious + likely deleterious missense variants at decreasing MAF cutoffs of 1%, 0.1%, 0.01% and singletons. For significant gene–cancer associations that appear in multiple MAF thresholds, the association with the highest significance was plotted. Only gene– cancer associations at M1 and M2 masks are labelled. **c,** Positive predictive value of known germline cancer-susceptibility gene panels (n=18) along with ExWAS-suggested panels (n=10) across cancer phenotypes for which LoF or deleterious missense variants were significantly enriched in participants with corresponding cancers. Numbers in x-axis represent number of cancer and non-cancer participants tested. The list of genes within each panel are provided in supplementary table 15. **d,** Percentage of cancer patients who might have adverse effect or non-response to cancer drugs reported in CGI database, grouped by cancer drug panels and variants considered. The bars are displayed in the descending order of proportion of patients carrying LoF or deleterious missense variants in the target genes within a panel. Numbers in x-axis represent number of cancer patients tested. **e,** PheWAS plot showing phenome-wide associations between ExWAS-suggested variants and gene burdens (n=66) and phenotypes (n=187) stratified by 14 phenotype groups. The red dashed line represents the significance threshold at P=0.001 to account for false discovery rate. Only non-neoplastic phenotypes from significant associations are labelled.

Genes representing 62% (24 of 39) of the associations did not have previously known relationships with cancer in the OMIM, GWAS catalogue, AstraZeneca PheWAS (AZ-PheWAS), or FinnGen release 12 databases. In total, 21 gene–cancer relationships were identified across these databases: GWAS catalogue (n=10), AZ-PheWAS (n=7) and FinnGen (n=4) databases. However, all 10 GWAS catalogue associations represented non-coding or synonymous variants; In contrast, 9 missense or LoF variant associations were identified from AZ-PheWAS and FinnGen by applying a more lenient significance threshold of P≤1×10^-5^. Of these, two (ANKRD11, KRT81) passed with suggestive threshold of P≤1×10^-6^.

The subgroup analysis revealed that majority of cohort-wide significant LoF and deleterious variant associations were driven by either of the ancestry subgroup (Supplementary Table 9). However, two associations were observed specifically in the Pakistani subgroup and did not reach significance at the cohort level: a splice site variant in TRPM2 gene associated with lymphoma and a deleterious missense variant in METTL13 (Ser425Pro), a known oncogene, associated with solid cancer. Notably, CBR1:*p.*Arg74Pro was the only cohort-wide significant variant whose association with solid cancer was not replicated in any of the ancestry subgroups. This lack of replication was possibly due to the ultra-rare nature of the variant, which did not pass the stringent minor allele count filter of five in any of the subgroup.

### Gene-level variant aggregation and associations

We performed gene-level association analyses by aggregating the effect of qualifying protein-altering variants, employing the omnibus strategy SKAT-O that combines traditional gene burden test with variance component test (Methods). We applied 12 distinct sets of qualifying variant filters (masks) to test the association of 22 cancer phenotypes with 16,821 genes. The 12 masks represented the aggregation of (i) LoF, (ii) LoF + deleterious missense, (iii) LoF + deleterious and likely deleterious missense variants at MAF cutoffs of 1%, 0.1%, 0.01% and singletons.

Using a P value threshold of 6.2×10^-6^ (Methods), we identified 31 significant associations, involving 25 genes and 28 gene-cancer pairs. 94% of these associations were driven by the accumulation of rare variants (Figure 4b; Supplementary Table 10). The most significant association was observed for singleton LoF variants in *NF1*, which encodes the tumour suppressor protein neurofibromin, and was associated with adult solid cancers (P=1.4×10^-12^). Other genes where LoF variants were associated with increased cancer risk included ARHGAP45 (skin cancer); BRCA1 (breast cancer); FKBP6 (lung cancer); and ZNF155 (prostate cancer). In all cases, the directions of effect were positive, meaning the aggregation of variants increases cancer risk, with the exception of ARID2, a tumour suppressor gene, which was associated with a decreased risk of all cancer combined (P=1.9×10^-6^).

As with the variant level analysis, 64% (16 of 25) of the significant genes did not have previously reported associations with cancer in any of the public databases we investigated. Only two associations were directly supported by existing databases: BRCA1 gene with breast cancer (OMIM and AZ-PheWAS), and NF1 gene with overall cancer overall (AZ-PheWAS). For eight genes (HLA-DMB, LAMB2, NF1, PKP1, SEC16A, SKA2, TMPRSS15, ZBTB10) we extended their known cancer-associations; and found novel cancer associations for further 16 genes. The large proportion of previously un-replicated cancer associations underscores the novelty of our findings and provides valuable insights into the phenotypic consequences of protein-altering variants in the South Asian population, potentially revealing new therapeutic targets.

Among the significant gene-level associations, only 25% (7 of 28) were also detectable via variant-level analysis. This highlights that aggregated analyses can identify rare variant associations that are undetectable through single-variant-based approaches, particularly for singleton and ultrarare variants. This enhanced detection was further supported by the subgroup analysis (Supplementary Table 11), where several associations were significant only within specific ancestry. When aggregating LoF and/or deleterious missense variants, approximately 70% of the significant associations were specific to one ancestry group (11 out of 16 in Bangladeshi subgroup; 7 out of 10 in Pakistani) and did not reach cohort-wide significance. On the contrary, six (out of 13) cohort-wide significant associations did not reach significance at the individual ancestry level.

### Pleiotropy and functional context

We conducted a phenome-wide association study (PheWAS) for the 66 ExWAS-derived unique variants and gene burdens. Using a study-specific collection of 215 phenotypes, we interrogated 187 phenotypes for associations observed in at least 50 participants (Methods). After correcting for false discovery rate, we identified 270 significant phenome-wide associations at P≤0.001 (Figure 4e; Supplementary Table 12). These associations were predominantly concentrated on neoplasm-specific phenotypes (n=183; 68%), representing a two-fold increase over the 78 exome-wide associations observed. About 20% of the additional neoplasm-specific associations (21 out of 105) were linked to low-prevalence cancers, such as stomach cancer or CNS cancers.

Among the non-neoplastic phenotypes, digestive system diseases had the highest proportion of pleiotropic associations with cancer-susceptibility genes (21%). Comorbidity analysis revealed a significant association between gastrointestinal polyp and digestive cancers, likely driven by LoF variants in NF1 gene which may serve as a useful marker for screening. Other notable pleiotropic associations included ulcerative colitis and colorectal cancer (PDC:p.Glu85Asp), benign CNS tumour and endocrine cancers (H1-1:*p*.Lys26Asn), and end-stage renal disease and urinary cancer (CLEC16A:*p*.Gly422Val). Additionally, we identified a pleiotropic effect of *FKBP6* gene, which was associated with both lung cancer and various mental disorders (ADHD, bipolar disorder, schizophrenia and psychosis). Although the involvement of other FKBP family proteins in lung cancer and mental disorders has been discussed in literature,^36,37^ FKBP6 itself has not been strongly linked to these conditions.

Among the genotypes, CLEC16A:*p.*Gly422Val, a possibly deleterious missense variant, had the highest number of pleiotropic associations (n=16), followed by RNF111:*p.*His355Tyr, a deleterious missense variant, with 10 associations. Although prior reports have linked these genes or their variants to other phenotypes (Supplementary Tables 8,10), 93% of the associations were not documented in any of the public databases queried. For example, 114 individuals (0.26%) carried at least one LoF variants in the *BRCA1* gene, with an increased risk of cancers previously linked to *BRCA1* mutations, including breast cancer (log-OR: 2.4 [1.6-3.3]; P=6.5×10^−9^) and ovarian cancer (log-OR: 3.1[2.0-4.1]; P=9.6×10^-9^)^38^ Interestingly, these individuals also exhibited an increased risk of Barretts oesophagus (log-OR: 2.4 [1.2-3.6]; P=7.6×10^-5^), a known precursor to oesophageal cancer. This suggests an under-recognized link between pathogenic *BRCA1* mutations and oesophageal cancer,^39^ especially in the context of familial neoplasms. This example highlights how our approach can uncover a broad spectrum of ancestry-specific phenotypes associated with protein-coding variations.

### Diagnostic and therapeutic Implications

We evaluated the efficacy of current cancer-susceptibility gene panels in SAS-BP population and compared them with panels derived from our study. The analysis focused on 295 unique genes across 17 cancer phenotypes: 96 genes tested clinically, as suggested by the NHS Genomic Test Directory for Cancer v11 (NGTDC);^40^ 196 genes conferring susceptibility to cancer, as reported in the Genomics England PanelApp;^41^ and 54 genes significantly associated with cancer (variants collapsed at gene level) from our study (G&H).

When assessing these panels for the presence of LoF and deleterious missense variants, we identified significant enrichment of variants in 28 panels across 10 cancer types, with PPV ranging from 0.6% to 21.4% (Fig. 4c, Supplementary Table 13). Notably, G&H-derived panels demonstrated the highest PPV for 5 out of 10 cancers, including 21.4% for *solid* cancers using LoF variants, and were the only panels significantly associated with bladder and skin cancer. The overall low PPV, with only 6 out of 124 panels exceeding 5% PPV, highlights the ineffectiveness of current cancer gene panels, which were primarily developed based on evidence from European/White populations, in minority ancestry groups. This underscores the need for either ancestry specific panels or expansion of existing panels to incorporate pan-ancestry data.

A similar challenge was observed in pharmacogenomics, where we examined the potential for tailoring cancer treatments in SAS-BP cancer patients based on their germline genomics. We found that the differentially mutated germline genes in our cohort were not significantly enriched for drug biomarkers in either a cancer-specific biomarker database (Cancer Genome Interpreter, CGI) or a broader gene–drug– disease interaction database (Pharmacogenomics Knowledgebase, PharmGKB) (Supplementary Fig. 6).^42,43^ Out of the 58 genes examined, only 2 were present in the 120 genes curated in the CGI database and 5 were included in the 1,468 for PharmGKB genes.

While somatic variants are increasingly targeted in therapeutic interventions, germline variants also play a critical role in predicting adverse drug effects or drug responses. To address this, we interrogated the CGI database to estimate the proportion of SAS-BP cancer patients who might experience adverse effects or non-response to cancer drugs based on their germline genomics. Among patients with LoF or deleterious variants, which are key targets in cancer pharmacogenomics, we found that up to 7% of total cancer patients could be impacted, with respiratory cancers showing the maximum impact (up to 10%). Focusing exclusively on the LoF variants, we found that current drugs could negatively affect 1% patients (Fig. 4d).

### Cross-ancestry validation of genetic findings

We aimed to evaluate the consistency and divergence of germline genetic signals across cancer patients of SAS-BP, SAS-nBP, and European ancestries. Whole-genome sequencing data from Genomics England Cancer cohort (n = 135 SAS-BP; 404 SAS-nBP; 12,980 European) were used in this analysis, focusing on coding regions. We focused our evaluation to a non-redundant set of 39 variants and 26 gene-level aggregates previously identified as germline susceptibility signals in SAS-BP individuals (Supplementary Tables 8, 10). Logistic regression models were applied to assess whether these candidate variants and genes exhibited differential mutation patterns in European or SAS-nBP cancer patients. Due to the limited sample size of the SAS-BP subgroup, analyses were performed for all cancers combined (n = 135), solid tumours (n = 125), and breast cancer (n = 45).

At the variant level, only two candidates passed the minor allele count threshold (MAC ≥ 3), but neither showed nominal significance (P≤0.05). Gene-level aggregates were defined using three aggregation models: (M1) loss-of-function (LoF) variants, (M2) LoF plus deleterious missense variants, and (M3) LoF plus deleterious or possibly deleterious missense variants. Among 78 gene-cancer comparisons (26 genes × 3 cancers), 19 (24%) demonstrated nominally significant associations (P≤0.05) when comparing SAS-BP and European cohorts. Importantly, all significant associations were in positive direction, suggesting that the implicated genes represent germline cancer susceptibility signals enriched in South Asian populations.

When comparing SAS-BP to SAS-nBP, six associations reached nominal significance. These included FAM166C (M3), SEC16A (M2), and TMPRSS15 (M3), all of which also showed reduced mutational burden in Europeans. This indicates that these germline susceptibility loci may be specifically enriched among Bangladeshi and Pakistani cancer patients, further highlighting the need for ancestry-tailored approaches in cancer.

## Discussion

This study provides a comprehensive genome-phenome analysis of cancer in a British South Asian population, leveraging the largest clinical history-linked exome-sequenced cohort of individuals of Bangladeshi and Pakistani ancestry. By integrating electronic health records, clinical phenotypes, and whole-exome sequencing, we identified novel genetic and clinical determinants of cancer risk in this underrepresented ethnic group. Our findings demonstrate that current cancer diagnostic and therapeutic paradigms, which are primarily based on European populations, may not be fully applicable to South Asian populations, underscoring the need for population-specific approached in cancer care.

### However, our work is subject to several limitations

First, the lack of external validation of our results stems, in part, from the scarcity of non-European studies that combine genetic with health record data, particularly for South Asians. The case numbers in existing cohorts such as the UK Biobank or 100K GP were insufficient for meaningful replication at the level of primary cancer types.

Second, despite efforts to assemble a large and representative cohort, statistical power remains limited for certain cancers due to small sample sizes. Furthermore, the heterogeneity within South Asian diaspora — encompassing individuals of diverse regional, linguistic, religious, and socio-economic backgrounds — may limit the generalizability of our findings to other South Asian subgroups, notably individuals of Indian descent.

Third, while ExWAS is a powerful tool for identifying genetic variants in coding regions associated with disease risk, it captures only a fraction of the heritable risk and overlooks structural variants and epigenetic factors. Additionally, the cross-sectional nature of EHR data precludes causal inference regarding the temporal relationship between risk factors and cancer outcomes. However, the integration of pleiotropy, comorbidity, and drug-target analyses offers functional context to many of our findings.

A key strength of our study is the enhanced accuracy of phenotype definitions through the integration of both primary and secondary care records. Unlike many phenome-wide genetic studies that rely solely on ICD-based billing codes from secondary care— an approach prone to phenotype misclassification^27^— our approach developed a more robust phecode framework by incorporating structured primary care data. This comprehensive coding strategy improves phenotypic precision and adds an additional layer of confidence to our association findings.

Our results highlight several important clinical insights. Notably, we observe earlier age of cancer onset in South Asian individuals, particularly among Bangladeshi participants, compared to national benchmarks. This finding underscores the need to re-evaluate current age-based screening guidelines for cancers such as breast and colorectal, which may not account for earlier disease onset in this population. We also identify 147 significant cancer-phenotype associations across various organ systems, with both systemic and organ-specific comorbidities playing a prominent role in cancer predisposition. Many associations—such as the link between fibromyalgia and haematological cancers, or gastrointestinal polyp and colorectal cancer—may point to diagnostic overlap, while others like hypertension and obesity, reinforce known modifiable risk factors that warrant targeted public health intervention in these communities.

Exome-wide analyses identified 39 rare coding variants and 31 gene-level associations with cancer phenotypes, over 60% of which had not been previously linked to cancer in major genomic databases. High-confidence findings such as *ZNF155* in prostate cancer and *NF1* in adult solid tumours, provide new insights into cancer susceptibility, alongside pleiotropic variants implicated in both cancer and non-neoplastic diseases. Importantly, many of these associations were ancestry-specific and undetectable at broader cohort analyses, highlighting the relevance of substructure-aware analyses in genetically clustered populations.

Current cancer susceptibility panels and pharmacogenomics resources, which are predominantly developed from cohorts of European ancestry, demonstrated limited predictive value in this South Asian population. Several high-penetrance gene–cancer associations identified in this study are absent from clinical testing panels, emphasizing the urgent need to diversify genomic resources and adapt cancer screening and treatment frameworks to better serve underrepresented populations.

In conclusion, our work provides foundational insights into the genomic and clinical landscape of cancer in British South Asian Bangladeshi and Pakistani populations, with significant implications for ancestry-specific diagnostics, risk stratification, and therapeutic targeting. This study lays the groundwork for the inclusion of diverse populations in future cancer genomics and precision oncology efforts to ensure equitable healthcare outcomes.

## Methods

### Study population

Genes & Health is a long-term, community-based study focused on British Pakistani and British Bangladeshi individuals aged 16 years and older living in the UK.^28^ At recruitment, participants provide a saliva sample for germline genotyping and/or molecular sequencing, complete a short questionnaire on basic demographic information and consent to linkage with local primary care, secondary care and national level EHRs. Since recruitment began in 2015, over 60,000 participants have been recruited. Ethical approval for the study was provided by the Southeast London National Research Ethics Committee (14/LO/1240). Data are available to researchers within a dedicated trusted research environment (TRE). An individual application to support data access for this study was granted by G&H Executive Board (reference: S00087).

### EHR data processing and curation

An initial cohort of 57,849 individuals with linked EHR data was curated from the December 2023 release of the dataset. Participant data were harmonised across primary and secondary care coding systems, including SNOMED (Systematized Nomenclature of Medicine), ICD10 (International Classification of Diseases, Tenth Revision), where applicable. The data were then organised into 216 binary phenotypes including 38 cancer phenotypes. Each phenotype was defined by a set of ICD-10 and SNOMED codes. Participants were assigned to a phenotype if at least one corresponding clinical code was recorded in any of the four EHR sources: Discovery East London data service (East London primary care), Barts Health NHS Trust (hospital records), Bradford Teaching Hospitals NHS Foundation Trust (hospital records) and NHS Digital (Hospital Episode Statistics and Cancer Registry). The earliest recorded date of diagnosis for each phenotype was used as the diagnosis date. Sex and ethnicity data were cross-referenced with participant-reported information from the questionnaire.

### Genetic ancestry correction of ethnicity

The genome-wide genotyping data were subject to rigorous quality control procedures, as outlined by Heng et al.^34^ Genotyping was performed using the Illumina Global Screening Array (GSAv3EAMD, build hg38) platform, and genetic ancestry (gAncestry) was inferred for each participant. Of the 51,166 genotypes individuals, 51,104 were identified as having either Bangladeshi or Pakistani gAncestry. Of the remaining 6,745 individuals with ambiguous gAncestry or missing genotyping data, 6,312 had self-reported ethnicity recorded as either Bangladeshi or Pakistani. In total, 57,416 individuals of only Bangladeshi or Pakistani origin were included in the analysis cohort for the comorbidity association study.

### Comorbidity association analyses

We performed associations analyses between cancer and non-cancer (control) phenotypes. For each cancer–non-cancer phenotype pair, a logistic regression model was used to estimate the odds of a cancer diagnosis in participants with pre-existing medical conditions (non-cancer phenotype) compared to those without the condition. Covariates included index age, index age squared, sex, and gAncestry-corrected ethnicity. For cancer cases, the index date was defined as the diagnosis date, while the recruitment date was used as the index date for control participants. A participant was classified as having a pre-existing condition if the earliest record of the condition occurred on or before the index date.

Controls for each model were selected under the following criteria: i) no prior cancer diagnoses; ii) exclusion of the opposite sex for analyses involving sex-specific phenotypes. Only phenotypes with a minimum of 100 cases were included in the analysis, resulting in the evaluation of 23 cancer phenotypes and 151 non-cancer phenotypes across 2,782 cancer cases and 54,634 controls. Subgroup analyses were conducted based on sex (male or female) and gAncestry-corrected ethnic subgroups (Bangladeshi or Pakistani), guided by the presence of significant interaction through likelihood ratio-based interaction test (P≤0.05). Bonferroni corrections were applied to adjust for multiple comparisons, and statistical significance was considered at P≤0.05.

### Whole exome sequencing

Exome sequencing data, linked with inferred gAncestry information, were available for 44,028 participants. These data were provided in analysis-ready files in both variant call format (VCF) and PGEN format. In summary, high-depth exome sequencing was performed using the Illumina Novaseq 6000 platform (150bp paired-end) with the Twist Clinical Research Alliance reagent at Broad Institute (Standard Germline Exome v6). Mapping to hg38 reference genome was performed with the BWA MEM algorithm. Variants were called using the GATK HaplotypeCaller (Y chromosome genotypes were not called). The quality control (QC) process excluded samples failing sample QC, as well as variants failing a set of stringent filters (QD < 2.0, FS > 60, MQ < 30.0) and genotype quality metrics (call rate > 95%, depth > 10, quality score > 20). Due to issues with ploidy when using HaplotypeCaller, variants on chromosome X were excluded from the analysis-ready files. After removing individuals with ambiguous gAncestry or missing genotyping data, the analysis cohort was restricted to 43,462 individuals with inferred Bangladeshi or Pakistani gAncestry for subsequent ExWAS and downstream analyses.

### Variant annotation

Variants were annotated with Ensembl’s Variant Effect Predictor (VEP) v.105.^44^ For each variant within a given gene, the consequence on the primary (canonical) transcript was extracted for subsequent analyses. Variants located outside the coding regions of the primary transcript, as well as monomorphic coding variants, were excluded. LoF variants were defined as those annotated as stop gained, start lost, splice donor, splice acceptor, stop lost or frameshift, provided the allele of interest was not the ancestral allele. To assess the deleteriousness of missense variants: five annotation tools were used: SIFT;^45^ PolyPhen HDIV and PolyPhen2 HVAR;^46^ LRT;^47^ and MutationTaster.^48^ Missense variants were classified as *likely deleterious* if predicted as deleterious by all five tools, *possibly deleteri*ous if predicted by at least one tool and *likely benign* if not predicted to be deleterious by any tool. Subsequent analyses were restricted to LoF variants, likely deleterious and possibly deleterious variants.

### Single-variant association analyses

Association analyses were performed using the two-step exome-wide regression approach implemented in REGENIE.^49^ In step 1, we included variants with MAF>1%, with less than 5% genotype or sample missingness, passing the Hardy–Weinberg equilibrium test (P>10^−15^) and linkage disequilibrium (LD) pruning (1,000 variant windows, 100 variant sliding windows and r^2^ <0.9). The choice of REGENIE as the genetic analyses tool was based on its ability to account for population structure and relatedness in step 1 through the estimation of a genome-wide polygenic effect component.

In step 2, logistic regression model with Firth correction (for associations reaching nominal significance; P≤0.01) was employed to test the association between each cancer type and rare or low-frequency variants (MAF≤1%). The regression models included leave-one-chromosome-out predictors obtained from step 1 as an offset, with the following covariates (i) index age, index age squared, sex, inferred gAncestry from genotyping array data; and (ii) 10 ancestry-informative principal components (PCs) derived from the analysis of LD-pruned (500 variant windows, 50 variant sliding windows and r2 < 0.2) common variants (MAF > 1%) from the exome data. We corrected for inferred gAncestry from array data to mitigate limitations of using PCs from common variants in correcting fine-scale population structure.^50,51^ Only variants with a minor allele count (MAC) of five or more were analysed. We defined significant associations by a P-value threshold of 3×10^−6^, corresponding to the nominal significance level at which one positive association would be expected from the maximum number of tests (n=331,291) performed for the cancer phenotype with the largest number of cases, i.e., all cancer combined (Supplementary Table 15).

### Gene-based collapsing association analyses

For gene-level association analyses, we aggregate low-frequency variants (MAF≤1%) within each gene that fit a predefined set of criteria – known as masks. Three categories of masks were considered for each gene: LoF variants (M1), LOF and likely deleterious missense variants (M2), and LOF and likely or possibly deleterious missense variants (M3). Each mask was further subdivided into four separate sub-masks based on the alternative allele frequency of the screened variants: MAF≤1%, MAF≤0.1%, MAF≤0.01%, and singletons only. Variants that met the criteria for each mask were collapsed into a single gene-level variable by taking the maximum number of minor alleles across the sites (MAX_MA) - the default strategy in REGENIE – which treats the presence of any rare variant in the target gene as a potential risk contributor. To address the limitations of traditional burden tests, which can lose power when variant effects differ in direction, we employed the SKAT-O (Optimal sequence kernel association test) method. This test combines a burden test and a variance component test, providing greater power when variant effects oppose each other, by computing weighted averages of each statistic.^52^

The above protocol for collapsing analyses was implemented in step 2 of REGENIE, using the same set of covariates as in the single-variant analyses. To address case-control imbalance, Firth correction was applied to calibrate the test statistics when the association reached nominal significance of P≤0.01. A total of up to 12 SKAT-O tests (3 masks x 4 sub-masks) were performed for each potential gene–cancer association (Supplementary Fig. 5), with only the most significant association at gene level reported. Significant associations were defined as those with P≤6.2×10^−6^, corresponding to the nominal threshold at which one positive association would be expected out of the maximum number of tests (n=161,671) performed for the cancer phenotype with the largest number of cases, i.e., all cancer combined (Supplementary Table 15).

### Ancestry specific signals

We conducted both variant-level and gene-based ExWAS for individual ancestry groups, following the same protocol outlined above. Individual cancer phenotypes with at least 50 cases were included in the analysis. As a result, ovarian and uterine cancers were excluded from the Bangladeshi subgroup, while cervical, endocrine, lung, non-melanoma skin, ovarian, respiratory and uterine cancers were excluded from the Pakistani subgroup. Separate P-value thresholds were applied for detecting significant associations in each ancestry subgroups, set at the level where one significant association would be expected from the maximum number of tests performed. For single variant tests, the thresholds were 4.3×10^−6^ (Bangladeshi) and 6.3×10^−6^ (Pakistani). For gene-based tests, these were 7.0×10^−6^ and 7.5×10^−6^ respectively.

### Benchmarking against external resources

We assessed the overlap between our significant variant/gene–cancer associations (collapsed at the gene level) and known gene-trait associations from four sources: i) the GWAS catalogue;^53^ ii) the OMIM database;^54^ iii) the AstraZeneca PheWAS Portal (470K WES data of European ancestry);^55^ and iii) the FinnGen release 12;^56^ all last accessed in June 2025. For GWAs catalogue and OMIM database, we considered any variants within a gene (or in nearby regulatory/intergenic regions) that had been reported as associated with any cancer phenotype, irrespective of the variant type. We also identified genes with reported associations to other non-cancer binary phenotypes. Associations from the AstraZeneca PheWAS Portal and FinnGen were considered if reported with P≤10^−5^. In addition, we checked South Asian ancestry specific associations in the AstraZeneca PheWAS Portal using 500K multi-ancestry whole genome sequencing data.

### Phenome-wide association study

We used a PheWAS approach to identify the phenotypes associated with unique gene masks or single variants that reached significance across cancer phenotypes in the previous steps. For each gene mask, a modified version of MAX_MA strategy was used to collapse qualifying variants into a binary entity representing presence of the minor allele across the sites. We constructed a study-specific disease phecode dictionary, comprising 215 phenotypes mapped from various ICD-10 and SNOMED codes. These phecodes were curated and assigned to participants as described above. For each phecode, participants with the respective phenotype were classified as cases, while all others were considered controls. Association between each phecode and each gene mask or single variant were tested using a logistic regression model, adjusting for the same covariates as in the ExWAS. Only phecodes with at least 50 cases were included, resulting in 187 total phecodes being analysed.

### Assessment of genomic test panel genes

We tested the efficacy of cancer-susceptibility gene panels recommended for germline testing in current clinical settings. The genes selected for analysis were derived from those included in the NHS National Genomic Test Directory for Cancer (NGTDC) v11,^40^ as well as those conferring cancer susceptibility and available for testing in the Genomics England PanelApp.^41^ The latter are rated according to a traffic light system (Green, Amber and Red) indicating high, moderate or low degree of evidence of association with cancer phenotypes. The final list included 244 unique genes across 17 cancer phenotypes in our study (Supplementary Table 16).

To evaluate the relationship between a gene panel and participants’ germline status, we used Fisher’s exact test. Genes were grouped into three panels: i) NGTDC; ii) genes with high evidence of association from Genomics England PanelApp Green list; iii) genes with moderate or low evidence of association in Genomics England PanelApp Amber or Red lists. A fourth panel, comprising significant genes and variants (mapped to genes) from our study was included for comparison (labelled G&H). Participants’ germline status was categorised into three groups reflecting the increasing mutational burden from the three gene masks in the ExWAS study. For each of the 17 cancer phenotypes, we created a contingency table to compare the number of cases with mutations in at least one susceptibility gene to the number of controls with the same criteria. Positive predictive values (PPV), odds ratios (OR) and P-values were reported.

### Enrichment of drug targets

We assessed the enrichment of drug targets in our cancer cohort using three publicly available drug target databases.

The first list was derived from the cancer-specific biomarker database CGI (last accessed on 5 June 2025)^42^, which reports the putative effects of mutations on cancer phenotypes as ‘Responsive’, ‘Resistant’, ‘No Response’ or ‘Increased Toxicity’. This list includes drug targets for various cancers suggested by Food and Drug Administration (FDA) guidelines (n=96), National Comprehensive Cancer Network (NCCN) guidelines (n=34), and those reported in clinical trials (n=192), case reports (n=148) and pre-clinical studies (n=260). For each unique (cancer x drug family x effect) tuple, we collapsed the entry at gene level resulting in a final list of 874 entries, spanning 27 cancer types, 134 genes, and 139 drug families (Supplementary Table 17).

The second list was derived from the PharmGKB,^43^ which curates potential gene–drug and gene–disease interactions, through an extensive review of sources such as the FDA biomarker list, the Clinical Pharmacogenetics Implementation Consortium (CPIC) genes–drugs interaction list, and published literature. From the interaction table, we identified the genes which had verified associations with any drugs. Our final list included 1,643 unique genes targeted by 1292 drugs (Supplementary Table 18).

We assessed the relationship between drug targets and gene–cancer associations using Fisher’s exact test. Genes were divided into two sets: i)significantly associated single variants and gene masks mapped to the corresponding genes (n=58); ii) genes tested in the ExWAS with non-significant associations (n = 16,763). For each drug target list, we created a contingency table that compared the number of significant genes from our study intersecting with the drug targets against the number of non-significant genes that intersected with the list.

To estimate the proportion of patients who might experience adverse effects or non-responsiveness to therapeutic regimens, we examined drugs listed in the CGI panel reported with the effects ‘Resistant’, ‘No Response’ or ‘Increased Toxicity’. We identified patients within each cancer group who carried at least one qualifying variant in any of the target genes in the respective panel. Variants were considered qualifying if they belonged to one of the three categories: LoF, deleterious and possibly deleterious, irrespective of the allele frequency in the cohort.

### Processing of the Genomics England dataset

The GEL 100K GP release v19 contains whole genome germline-sequenced data for participants with cancer and rare diseases, linked to their associated clinical data, and is available to researchers within a dedicated TRE. Participants were assigned to one of the six ancestry supergroups - European, African, South Asian, East Asian, American Admix and Admix, inferred from genotyping data, using AggV2 - a multi-sample genomic VCF file made from release 10 of the 100K GP project and comprises 78,195 germline genomes aligned to GRCh38 and passed quality control.^57^

For a cross-ancestry validation study, we identified 23 cancer cohorts aligned to G&H cancer phenotypes, based on primary cancer type diagnoses from the GEL core cancer dataset. In addition, NHS England secondary dataset within GEL was interrogated for matched ICD-10 codes of 38 cancer phenotypes (see EHR data processing and curation section). For sex-specific cancers, the respective cohort was subsequently filtered to include only male or female participants as appropriate. For a more granular analysis of the South Asian population, participants were further categorised into two groups: i) individuals identified as having Bangladeshi or Pakistani ethnicity from the Participant Summary secondary dataset (SAS-BP); ii) the remaining individuals not of Bangladeshi or Pakistani ethnicity (SAS-nBP). This led to a validation cohort of 13,519 participants representing three groups: SAS-BP (n=135), SAS-nBP (n=404), and European (EUR; n=12,980).

The AggV2 file was used to generate summary statistics for samples and variants annotated by VEP (v109). To compare the germline profile of SAS-BP cancer patients with respective SAS-nBP and EUR cancer patients, we performed both variant- and gene-level association analyses. Gene level analysis was conducted by aggregating low-frequency coding region variants (MAF ≤ 1%) according to specific masks, outlined above. Individual germline genomes were passed through the GEL small variant workflow (v3.1.4) to extract the coding region variants within specific genes. Briefly, this takes the raw germline VCF files, filters on the participants and the relevant genes, annotates the variants, applies the masks, and generates a single multi-sample VCF file. For each cancer phenotype and comparison groups (SAS-BP vs SAS-nBP, SAS-BP vs EUR), individual logistic regression models were employed to test the association with either a single variant- or gene-level variable. The gene-level variable, defined by the given gene-mask combination, was assigned the value of maximum number of minor alleles across the qualifying sites (MAX_MA strategy). The regression models included sex as covariate for cancers not specific to any sex.

## Data availability

Clinical and molecular data from Genes & Health is available through the Genes and Health Research Environment via application at https://www.genesandhealth.org/researchers/apply-for-access/. Applicants are required to fill in a Data Access Agreement, complete mandatory information governance training and pay a small fee for access to the trusted research environment to examine individual-level data. Only aggregate-level data can be exported from the trusted research environments for research purposes with restrictions to prevent the identification of individual participants through data linkage.

Genomic and phenotypic data for the 100K GP study participants are available through the Genomics England Research Environment via application at https://www.genomicsengland.co.uk/join-us: approval to access the anonymised data through the Genomics England Trusted Research Environment requires a research project proposal, and mandatory training on information governance.

## Code availability

Most of the data curation and downstream analyses were conducted using Python version 3.9.2. Variant level statistics were generated with PLINK version 2.0.0. REGENIE version 3.2.6. was used to conduct the ExWAS analyses. The PheWAS analyses were completed using R version 4.3.3 PheWAS package. Codes for all the analyses from the manuscript will be made available to researchers on application to the corresponding author via a release from a GitHub repository, under a Creative Commons Attribution-NonCommercial 4.0 licence (CC BY-NC 4.0; https://creativecommons.org/licenses/by-nc/4.0/), once access to the datasets in Genes and Health and the Genomics England Research Network is confirmed. As most of the codebase work on individual-level data, they need to be imported into the relevant trusted research environments to perform the analyses, with the results exported from the TREs subject to the individual TRE restrictions.

## Author Contributions

Methodology: ADU, CC; Formal Analysis: ADU, AJ, GJT, LGEJ; Data access and Curation: ADU, CC, G&H, AJ, GJT, LGEJ; Visualisation: ADU; Writing – original draft: ADU; Writing – revisions: ADU, CC; Writing – review & editing: all authors; Funding acquisition: CC; Supervision and Conceptualisation: ADU, CC.

All authors read and approved the final manuscript.

## Acknowledgments

This work was directly supported by Barts Charity (MGU0504) to C.C. This research was made possible through access to data in the Genes & Health study, which is core-funded by Wellcome (WT102627, WT210561), the Medical Research Council (UK) (M009017, MR/X009777/1, MR/X009920/1), Higher Education Funding Council for England Catalyst, Barts Charity (845/1796), Health Data Research UK (for London substantive site), and research delivery support from the NHS National Institute for Health Research Clinical Research Network (North Thames). We acknowledge the support of the National Institute for Health and Care Research Barts Biomedical Research Centre (NIHR203330); a delivery partnership of Barts Health NHS Trust, Queen Mary University of London, St George’s University Hospitals NHS Foundation Trust and St George’s University of London. Genes & Health has recently been funded by Alnylam Pharmaceuticals, Genomics PLC; and a Life Sciences Industry Consortium of AstraZeneca PLC, Bristol-Myers Squibb Company, GlaxoSmithKline Research and Development Limited, Maze Therapeutics Inc, Merck Sharp & Dohme LLC, Novo Nordisk A/S, Pfizer Inc, Takeda Development Centre Americas Inc. We thank Social Action for Health, Centre of The Cell, members of our Community Advisory Group, and staff who have recruited and collected data from volunteers. We thank the NIHR National Biosample Centre (UK Biocentre), the Social Genetic & Developmental Psychiatry Centre (King’s College London), Wellcome Sanger Institute, and Broad Institute for sample processing, genotyping, sequencing and variant annotation. This work uses data provided by patients and collected by the NHS as part of their care and support. This research utilised Queen Mary University of London’s Apocrita HPC facility, supported by QMUL Research-IT, http://doi.org/10.5281/zenodo.438045. We thank: Barts Health NHS Trust, NHS Clinical Commissioning Groups (City and Hackney, Waltham Forest, Tower Hamlets, Newham, Redbridge, Havering, Barking and Dagenham), East London NHS Foundation Trust, Bradford Teaching Hospitals NHS Foundation Trust, Public Health England (especially David Wyllie), Discovery Data Service/Endeavour Health Charitable Trust (especially David Stables), Voror Health Technologies Ltd (especially Sophie Don), NHS England (for what was NHS Digital) - for GDPR-compliant data sharing backed by individual written informed consent. Most of all we thank all of the volunteers participating in Genes & Health. A favourable ethical opinion for the main Genes & Health research study was granted by NRES Committee London - South East (reference 14/LO/1240) on 16 Sept 2014. Queen Mary University of London is the Sponsor, and Data Controller.

This research was also made possible through access to data in the National Genomic Research Library, the National Genomic Research Library, which is managed by Genomics England Limited (a wholly owned company of the Department of Health and Social Care). The National Genomic Research Library holds data provided by patients and collected by the NHS as part of their care and data collected as part of their participation in research. The National Genomic Research Library is funded by the National Institute for Health Research and NHS England. The Wellcome Trust, Cancer Research UK and the Medical Research Council have also funded research infrastructure.

